# Absolute quantification of brain deuterium metabolic imaging in healthy volunteers and glioblastoma patients at 7T

**DOI:** 10.1101/2025.10.08.25337571

**Authors:** Jabrane Karkouri, Masha Novoselova, Sarah Miller, Minghao Zhang, Alexandra Constantinou, Carina Graf, Daniel Atkinson, Brandon Tramm, Scott Schillak, Richard Mair, Kevin Brindle, Christopher T. Rodgers

## Abstract

**Purpose:** We present a method for absolute quantification of deuterated metabolites *in vivo* at 7T. We describe acquisition protocols and an analysis pipeline compatible with 7T Terra MRIs, and apply these using a ^2^H/^1^H receive array in healthy volunteers and glioblastoma patients.

**Methods:** B_1_^+^/B_1_^−^ maps from multiple CSI scans in a uniform phantom were used to derive calibrated coil weights for absolute quantification, validated in phantoms with known deuterated compounds. ^2^H MRSI was performed in twelve healthy volunteers (two post-[6,6-^2^H_2_]-glucose) and five glioblastoma patients (all post-[6,6-^2^H_2_]-glucose). Spectra were fitted with OXSA, and Glx/Lac compared between tumor and normal-appearing brain using linear mixed-effects models.

**Results:** Measured B_1_^+^ maps were 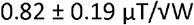 across the whole phantom. Natural abundance deuterium in water was 8.96 ± 0.7 mmol/L. Absolute maps of HDO, Glc, Glx, and Lac were acquired following [6,6-^2^H_2_]glucose. Rate maps showed higher Lac production in tumors (2.3 μmol/L/min, SE = 0.87) compared with normal-appearing regions (1.0 μmol/L/min, SE = 0.36; *p* < 0.01) and healthy brain (0.5 μmol/L/min, SE = 0.17; *p* < 0.01). Glx production was lower in tumors (3.8 μmol/L/min, SE = 0.44) relative to normal-appearing regions (6.0 μmol/L/min, SE = 0.36; *p* < 0.001) and healthy brain (9.2 μmol/L/min, SE = 0.61; *p* < 0.001).

**Conclusion:** We demonstrate robust absolute quantification for human 7T DMI. Glioblastomas showed elevated Lac and reduced Glx labeling relative to normal brain, with inter-patient heterogeneity consistent with an existence of different metabolic subtypes.

## 1 Introduction

Altered brain energy metabolism is a hallmark of prevalent diseases, including neurodegenerative disorders and cancer(1-7). Insufficient cellular energy impairs neuronal function, leading to cell death during ageing and as a result of neurodegenerative disease. Therapies aimed at restoring brain energy metabolism—such as oral supplementation with NAD^+^ or exogenous ketones, or intermittent fasting—are actively being investigated to treat Parkinson’s disease and other disorders(8-11). Tumors, by contrast, often display fundamentally different metabolism, with a preference for glycolysis(7,12-15). Non-invasive characterization of such metabolic changes could enable personalized treatment selection(16). Alternatively, tumor metabolism itself is an attractive therapeutic target; recent reviews highlight glycolysis, glutaminolysis, and mitochondrial pathways as key precision oncology targets (13,17,18).

Glioblastoma, the most aggressive brain cancer, is metabolically heterogeneous. Spatial transcriptomics and mass spectrometry imaging of ^13^C-glucose–infused patients reveal glycolytic and oxidative subtypes(19-22). These can coexist at the centimeter scale, suggesting they may be resolvable by clinical metabolic imaging. Because the glycolytic subtype is radiation-resistant and carries a poorer prognosis, imaging could inform prognosis and treatment selection. Clinical trials of energy-modifying therapies likewise require robust, non-invasive metabolic endpoints to permit trials in economically-viable cohorts.

Magnetic resonance spectroscopy (MRS) and spectroscopic imaging (MRSI) provide powerful *in vivo* tools to probe brain metabolism(23-26). A particularly promising emerging MRSI approach, called deuterium metabolic imaging (DMI), uses deuterium-labelled substrates to track metabolic fluxes in real time. DMI was introduced in 2018 at 11.7T(27) and has subsequently been applied *in vivo* by several groups(28-48).

DMI allows mapping of the partitioning of tissue glucose metabolism between glycolysis and mitochondrial oxidation. ^2^H-MRSI follows metabolism of [6,6-^2^H_2_]-glucose into glutamate/glutamine (Glx) via the TCA cycle and into lactate (Lac) via glycolysis, as pioneered by de Feyter and de Graaf(27,49-52). This provides unique insights into metabolic reprogramming and may serve as a radiation-free alternative to ^18^FDG-PET, as shown recently in Alzheimer’s disease(53).

Preclinical work supports DMI’s potential for monitoring treatment response(16,54-58), for grading tumours(59-61) and to determine subtypes of glioblastoma xenografts by quantifying [6,6-^2^H_2_]-glucose partitioning between Glx and Lac(62). Ultra-high-field MRI improves both SNR and also spectral separation (between HDO, Glc, Glx, and Lac), as demonstrated by studies at 9.4T(63) and 7T(64-71) compared to 3T(72-76).

Here, we focus on quantifying deuterium metabolites at 7T in healthy volunteers and glioblastoma patients. We adapted a method for *absolute quantification* (finding metabolite concentrations in mol/L) that was originally developed by *Purvis et al*.(77) for liver 7T ^31^P-MRS. We validated it in phantoms and in healthy volunteers without administration of a deuterated tracer. Then, we applied the method to quantify the temporal dynamics of HDO, Glc, Glx, and Lac concentrations after oral [6,6-^2^H_2_]glucose. We compared healthy tissue in volunteers with tumor-containing and normal-appearing regions in patients to reveal inter-patient and intra-tumor metabolic heterogeneity.

## 2 Theory

### 2.1 Quantitation framework

The key requirements for quantitative MRSI with a receive array are: (i) combining signals from array elements appropriately for quantitation, (ii) fitting spectral peaks, and (iii) correcting for metabolite and reference phantom relaxation.

We adapt “Method 1: Phantom Replacement” from *Purvis et al*. which was originally developed for 7T ^31^P-MRS of the human liver(77). This relies on acquiring data from a conductivity-matched phantom of known concentration and T_1_ with a matched protocol for calibration.

### 2.2 B_1_^+^mapping

The Phantom Replacement method requires knowledge of the coil’s B_1_^+^ and B_1_^−^ fields. Following Purvis *et al*.(77), we measured these using Variable Flip Angle (VFA) B_1_^+^ mapping on a uniform phantom(78). Multiple CSI scans were acquired with *N*_*V*_ =9 RF excitation voltages (*V1,V2*, ⃛ ). For each voxel, all the single-element spectra were concatenated to form a joint spectrum:

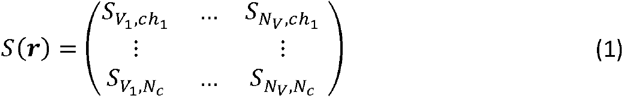

of size *N*_*c*_× (*N*_*V*_ *N*_*s*_ ), where *N*_*c*_ is the number of receive elements and *N*_*s*_ is the number of points in each spectrum. The noise covariance Ξ was estimated from a no-RF prescan. Whitened Singular Value Decomposition (WSVD) was then applied in each voxel to compute *N*_*c*_ × 1 coil weight vectors *w*(***r***) jointly for all the CSI scans, and maximum likelihood combined spectra *S*_*j*_ (***r***).

The combined spectrum with maximum signal was fitted in AMARES(79) to obtain a model spectrum *S*_*M*_ (***r***). Using linear least squares (“\” in MATLAB), complex single-element amplitudes *A*_*k,j*_(***r***) = *S*_*k,j*_(***r***)\ *S*_*M*_ (***r***) and combined amplitudes *A*_*j*_(***r***) = *S*_*j*_(***r***) \ *S*_*M*_ (***r***) were derived for each element *k* and voltage *j*.

Using the known T_R_ (1000ms) and phantom T_1,P_ (355ms), voxel-wise *B*_1_^+^maps were obtained by nonlinear least-squares fitting (lsqcurvefit, MATLAB) to:

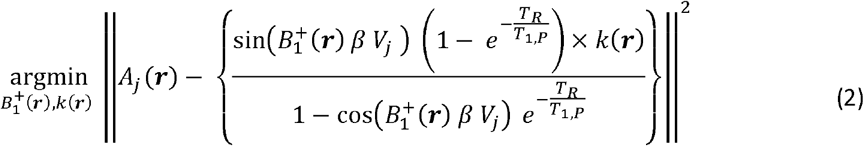

where 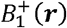 is the transmit field strength per volt in units of µT/V_RMS_, and 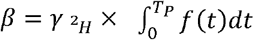 accounts for the deuterium gyromagnetic ratio 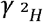 and the RF pulse shape *f*(*t*) integrated over the pulse duration *T*_*P*_,*T*_*R*_ is the repetition time, *T*_1,*P*_ the phantom *P*^2^H T_1_ and *k(****r***) is a scaling factor representing both coil receive sensitivity and the phantom concentration(80).

### 2.3 B_1_^-^ mapping

According to the Roemer signal model(81,82), the signal in the *k*^th^ element depends on the sample magnetisation and that element’s receive sensitivity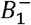. The magnetisation depends on the ^2^H concentration (known in the phantom), 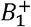and the pulse sequence. We include in “receive sensitivity” all fixed factors (cable losses, preamplifier/channel gains), which remain identical for reference and main scans. The overall phase of 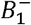 was defined by assuming the spectrum at the voltage giving maximum signal was real and positive.

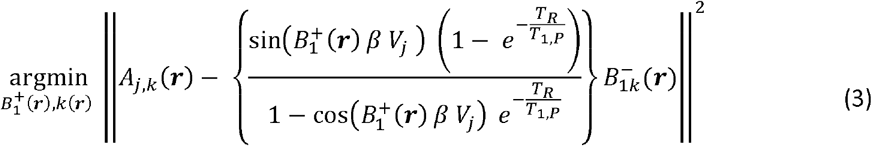

This was solved by linear least squares (“\” in MATLAB). Effectively, the scaling *k*(***r***) from the 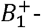mapping is distributed across receive elements, with relative phases captured since 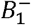is complex.

### 2.4 Uniform sensitivity coil combination

The joint WSVD weights *w*_*k*_ at position ***r*** are mathematically equivalent to Roemer’s uniform sensitivity weights (Eq. 27,(81)):

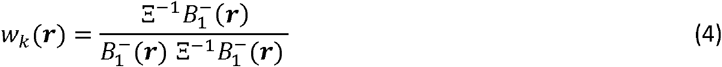

Roemer weights capture the complex scaling from spatially varying coil sensitivity, whereas WSVD— being data-driven—assumes an arbitrary overall sensitivity.

For quantitative analysis, we apply the reference-phantom-derived weights to combine per-element spectra from the main scan (e.g. an *in vivo* scan to be calibrated). This assumes that 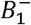 and noise covariance Ξ are similar between the reference and main scans, which holds if conductivity and coil loading are comparable. The combined spectra are then fitted with AMARES, giving metabolite 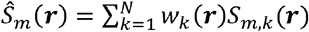 amplitudes for the main scan and 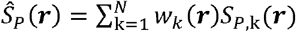 for the reference phantom.

Following Purvis *et al*. (Eq. 10), the metabolite concentration is:

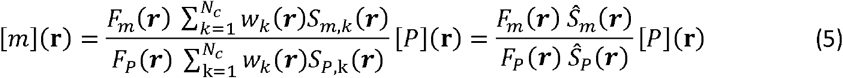

where *F*_*m*_ and *F*_*p*_ are saturation-correction factors for metabolite and the reference phantom, which has concentration [*P*], and *N*_*c*_ is the number of receive elements. For a spoiled CSI sequence:

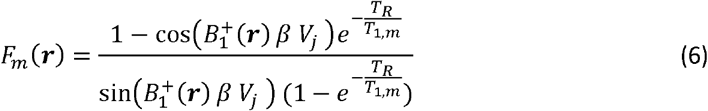

where *T*_*1,m*_ is the *T*_1_ of metabolite *m* (or phantom compound).

## 3 Methods

### 3.1 Hardware

All experiments were performed using a Magnetom Terra 7T MRI (Siemens, Erlangen, Germany) with a dual-tuned ^2^H/^1^H head coil (Virtumed LLC, Minneapolis, MN). Figure 1d shows a photograph of the coil, which comprises a 15-element Transverse Electromagnetic mode (TEM) resonator for ^2^H transmit (one rung is left out to allow visual stimulus), 16x ^2^H receive loops, and 4x ^1^H transmit/receive loops driven in quadrature.

**Figure 1:**
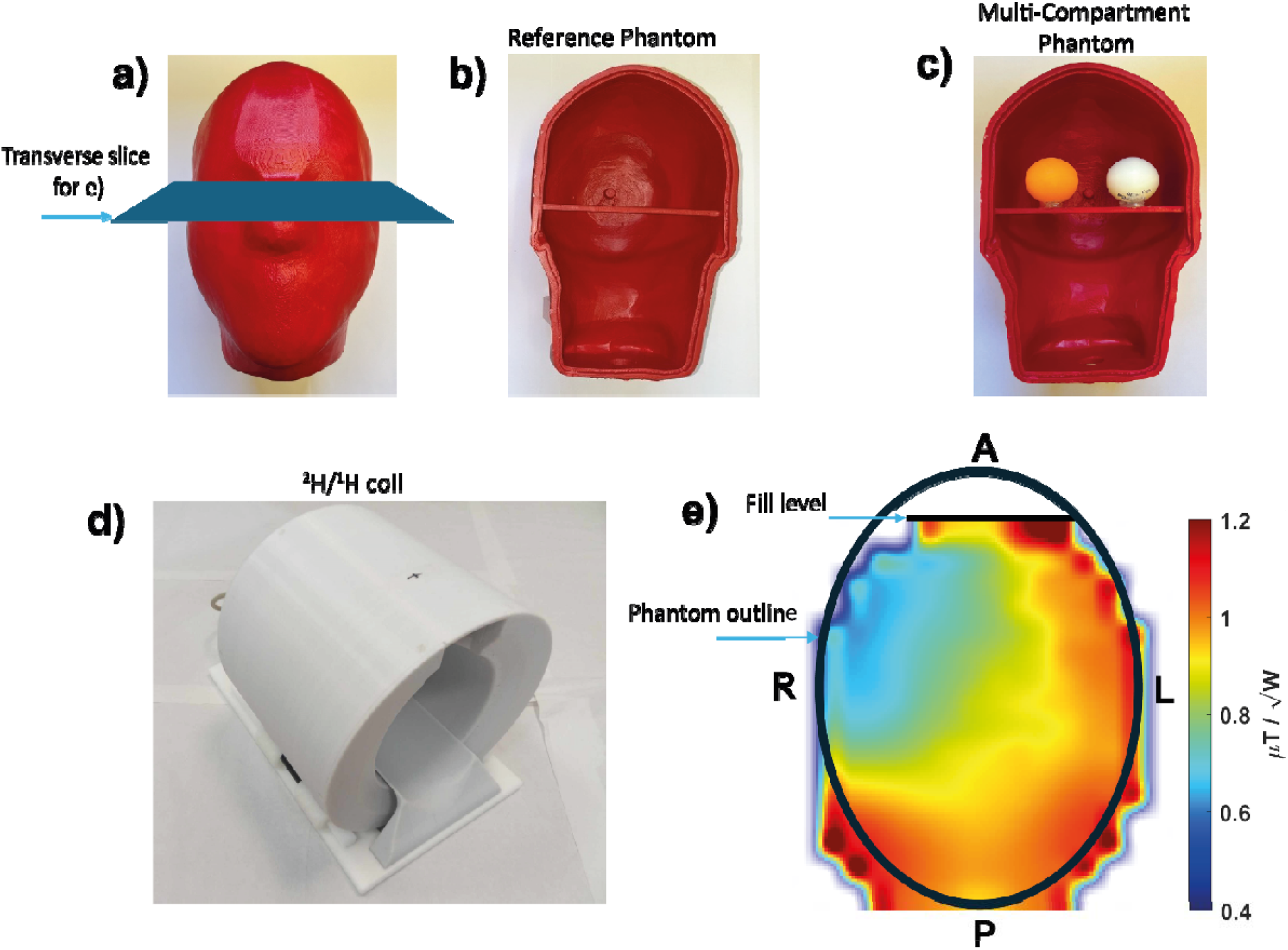
Custom 3D-printed head-shaped phantoms for calibration and validation. a) Photograph of the phantom housing. b) Reference phantom filled with deuterium oxide and water. c) Multi-compartment phantom containing two table tennis balls containing aqueous solutions of Lac and Fum, with the remaining space filled with deuterium oxide and water. d) Photograph of the dual-tuned 7 T ^2^H/^1^H head coil used for this study. e) B1+ map with the reference phantom (b) plotted for the mid-transverse slice illustrated in (a).

### 3.2 Reference phantom

We 3D-printed a custom head-shaped phantom (Figure 1a) based on the design by Shajan et al.(83), to which we added a threaded bottle neck and cap, and a support for the inner compartments (table tennis balls). It was printed from polylactic acid (80% infill, Bambu 3D printer, Shenzhen, China) and waterproofed with four coats of red rubber spray (PlastiDip UK Limited, Petersfield, UK). The phantom was filled with 4.5 L ultrapure water (Electrolube, Ashby de la Zouch, UK), 51 g deuterium oxide (yielding an HDO concentration of 1.14 mol/L), 0.25 g Dotarem (Guerbet, Princeton, USA), 20.93 g sodium chloride, and 8.4 g sodium benzoate. Chemicals were obtained from Sigma-Aldrich and used as supplied. Conductivity was estimated at 1.0 S/m(84), comparable to tissue values(85,86), where grey matter is 0.47 S/m, white matter 0.27 S/m and cerebrospinal fluid 2.04 S/m.

#### 3.3 Quantitation protocol

The scans need to implement this method are:

1. Main MRSI scan (*in vivo* or phantom) to be quantified.

2. Reference phantom B _1_^+^ maps to fix *w*_*k*_(***r***) in Eq. (4) and 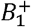 (***r***) in Eq. (3).

3. Phantom T_1_ determination.

4. Reference scan with matched protocol to determine *F*_*P*_(***r***) and *ŝ*_*P*_(***r***) in Eq. (5).

Data from steps 2-3 can be reused to quantify many MRSI scans providing their protocols match, including having identical position and orientation for their MRSI matrices.

### 3.4 B_1_^+^and B_1_^-^ maps

Reference phantom B_1_ ^+^ and B_1_ ^+^ maps were obtained by acquiring nine 3D CSI scans with 4ms rectangular excitation pulses having peak voltages ranging from 25V to 225V in 25V steps, 1 s TR, 16x16x8 matrix, 5 kHz spectral bandwidth, 1024 points, and acquisition weighting with 1 average at k=0. The total acquisition time was 1 hr 30 min.

### 3.5 Validation of absolute quantification

The reference phantom (Figure 1a-b) was scanned as part of the “Reference scan” with *T*_*R*_ = 250 ms, 16×16×8 matrix, 1 ms block excitation pulse at 210 V, 5 kHz spectral bandwidth over 1024 points, 6 averages, and acquisition weighting, giving a total time of 5 min. A second scan (i.e. the “main scan” for the purpose of this test) was performed on another day with identical parameters and grid position.

A second multi-compartment phantom was built using the same head-shaped shell but with two table-tennis balls glued inside (Figure 1c). Each contained 50 mL ultrapure water and 0.23 g sodium chloride. One was supplemented with [2,3-^2^H_2_]fumaric acid (CLM-1529; 0.99 g; MW 118.1 g/mol; 0.25 mol/L; deuterium concentration 0.5 mol/L) and the other with sodium L-[3,3,3-^2^H_2_]lactate (DLM-9071; 1 g; MW 115.03 g/mol; 0.26 mol/L; deuterium concentration 0.78 mol/L). Both were obtained from CK Isotopes Limited (Leicester, United Kingdom). The bulk head volume contained 9 g D_2_O and water, giving an HDO concentration of 0.2 mol/L.

To minimise voxel partial-volume effects, we used a 20×20×16 matrix while keeping other parameters identical, for a total acquisition time of 29 min 12 s. A matching calibration scan of the reference phantom was also acquired at this resolution. Following Purvis et al.(77), B_1_ maps were interpolated to this grid using MATLAB’s *imresize3*.*m* (bilinear interpolation).

#### 3.5.1 T_1_ measurements

Separate measurements were performed to determine the *T*_1_ values for each metabolite in each phantom, i.e. HDO in the reference phantom, and HDO, Lac, and Fum in the multi-compartment phantom.

The reference phantom was scanned alone, each table-tennis ball was scanned individually, and finally the complete multi-compartment phantom was scanned after assembly. The *T*_1_ values of each metabolite were measured using non-localized inversion recovery scans and fitted to a 3-parameter model. The inversion-recovery acquisitions had inversion times that were incremented from 25 ms to 8000 ms, 10 s TR, 1ms rectangular excitation pulses and 2ms rectangular inversion pulses both set to 150 V_rms_.

### 3.6 In vivo data acquisition

#### 3.6.1 Participants

Twelve healthy volunteers (6 females, 6 males, 29–58 years) and five glioblastoma patients (2 females, 3 males, 53–67 years) gave written consent. Ethics approvals were obtained from the University of Cambridge HBREC (HBREC.2021.08) and the London Camden & King’s Cross REC (21/PR/0828).

#### 3.6.2 Study protocol

Ten healthy volunteers were scanned using natural-abundance HDO only. The remaining two volunteers and all patients fasted ≥4 hours before receiving a glucose drink (0.75 g/kg body weight, max 60 g) of 6,6’-[^2^H_2_]glucose in ≈200 mL water prior to the 7T scan. Patients also underwent a 3T clinical scan during the same visit.

#### 3.6.3 7T MRI protocol

Participants were scanned lying head-first supine, with lights switched off at the main circuit breaker to reduce noise at the ^2^H frequency (Figure SI1). MRI acquisition began with ^1^H gradient-echo scouts (TE = 3.6 ms, TR = 8.6 ms, FOV = 400 mm^2^, 1.56 × 1.56 mm^2^ in-plane, 5 mm slice, 17° flip angle) followed by T1-weighted images (TE = 1.8 ms, TR = 1 s, FOV = 280 × 245 × 192 mm^3^, 1.09 × 1.09 mm^2^ in-plane, 1 mm slice, 5° flip angle).

B_0_ shimming was performed with the vendor’s “GRE-Brain” tool, and post-shim maps were acquired with dual-echo GRE (TE = 1.02/3.06 ms, TR = 18 ms, FOV = 256 × 256 × 256 mm^3^, 4 mm in-plane, 4 mm slice, 7° flip angle).

^2^H MRSI data were collected in blocks by repeating a UTE-CSI(87,88) sequence with 6.9 mL nominal resolution (16 × 16 × 8 matrix, FOV = 220 × 200 × 320 mm^3^, TR = 250 ms, 1 ms rectangular pulse at 210 V_RMS_, 5 kHz bandwidth), giving 4min59s per measurement. The CSI grid and parameters matched those of the phantom scans and were fixed across all participants.

#### 3.6.4 3T acquisition protocol

The five glioblastoma patients also underwent 3T imaging (Prisma, Siemens, Erlangen, Germany) using a standard brain protocol(89,90) including MPRAGE, T_2_ SPACE, T_2_-weighted turbo spin echo, and diffusion-weighted EPI, all following Gadovist contrast administration (Bayer New Zealand Limited, New Zealand).

### 3.7 Post-processing

Anatomical 3T images were co-registered to the 7T GRE scan using an affine transformation with 12-degrees of freedom and mutual information (imregtform, MATLAB). Data were processed in MATLAB R2022b with an updated OXSA toolbox supporting Siemens 7T Terra scanners, incorporating AMARES(91).

For ^2^H MRSI, analysis proceeded as follows (summarised in Figure 2):

**Figure 2:**
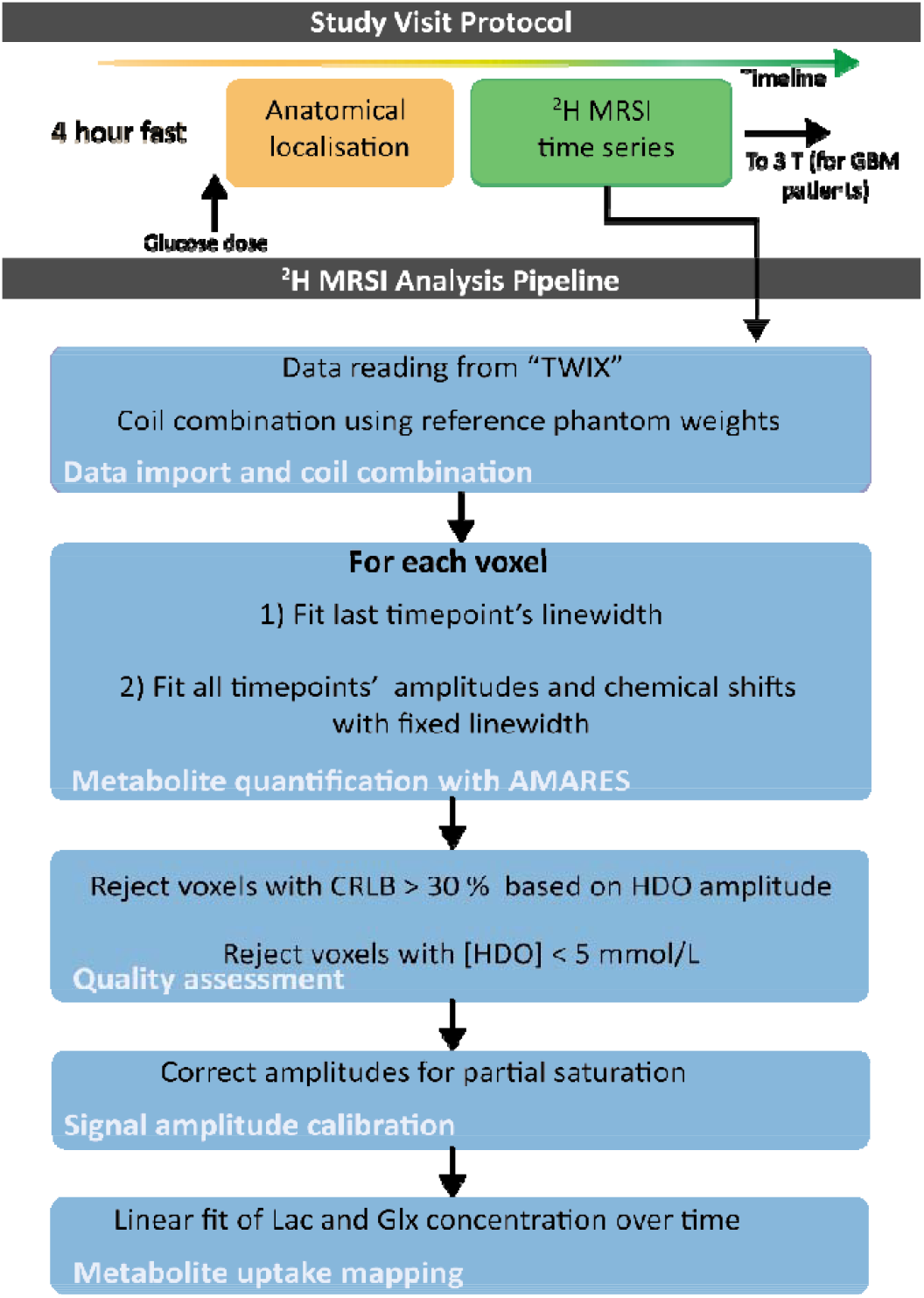
7T DMI protocol (top) and ^2^H MRSI processing steps (bottom). All participants fasted at least for 4h prior to the scan, which included the uptake of a deuterated glucose drink, anatomical localization and a series of 3D CSI scans over time. The data were then exported for further analysis. A metabolite mask was generated from the HDO peak amplitude and its CRLB values before amplitudes were corrected for partial saturation.

1. Raw *k*-space data (Siemens “TWIX” format) were Fourier transformed into image and frequency space.
2. For each voxel:
  a. Multi-channel signals were combined using Roemer weights calibrated on the reference phantom (see Section 2 above for details).
  b. The last timepoint was quantified using prior knowledge of 4 Lorentzian peaks.
  c. The prior knowledge was updated in memory to fix those linewidths before fitting data from all other timepoints.
  d. Amplitudes were corrected for partial saturation using literature T_1_ values (Lac: 297 ms; Glc: 66 ms; Glx: 149 ms; HDO: 450 ms)(28,44,92).

#### 3.7.1 Quality Control and Thresholding

Cramér-Rao Lower Bounds (CRLB) were computed to estimate uncertainty.

Mask #1 excluded voxels with [HDO]<5 mmol/L, or HDO concentration CRLB>30%.

Mask #2 was taken first from mask #1 and also excluded voxels with Lac/(Lac + Glx) concentration ratio CRLB > 30%.

#### 3.7.2 ROI analysis

To compare metabolism between healthy controls, tumor regions, and contralateral normal-appearing brain, three ROIs were defined In patients, separate ROIs were outlined based on 3T images: tumor (ROI_tumor_) and contralateral normal-appearing white matter (ROI_NAWM_). In glucose-ingesting healthy controls, a similar size ROI placed in the center of the brain was used (ROI_HV_).Voxel-wise data within each ROI were averaged to yield an ROI mean value per region, per patient, per time point. These ROI-mean data were used for all statistical analyses (see Figure SI2).

### 3.8 Time series analysis

Following the approach by Li *et al*.(93) and Niess *et al*.(41), to test whether there were statistically significant changes in metabolite concentrations over time, we fitted Lac and Glx concentrations to a linear model(41,71):

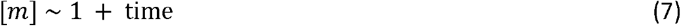

This produces a summary of the rate of increase of Lac, i.e. 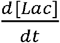, and Glx, i.e. 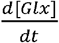, for the whole scan.

Maps of the product selectivity toward lactate 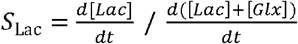 were then investigated with mask #2 (section 2.7.1).

### 3.9 Statistical analysis

#### 3.9.1 Regional influence on Glx and Lac temporal dynamics

To assess whether tumor or normal-appearing regions had different Glx and Lac kinetics, we used two complementary statistical approaches. First, we performed paired comparisons using the non-parametric Wilcoxon Signed-Rank Test(94), matching each tumor value with the corresponding value in normal-appearing brain from the same patient at the same time point.

Second, we applied stepwise linear modelling (MATLAB “stepwiselm.m”) with the Akaike Information Criterion (AIC) to identify predictors of Glx or Lac concentrations. Candidate predictors included Patient ID, tissue type (tumor vs normal-appearing), and nuisance variables (age, gender, time). The best-fitting models were then refined using linear mixed-effects modelling (MATLAB “fitlme.m”) to account for repeated measurements within patients (Model 1)(95).

#### 3.9.2 Inter-Patient Metabolic Heterogeneity in Tumour Regions

To examine inter-patient metabolic variability, analysis was restricted to tumor regions and two linear mixed-effects models were fitted. Model 2:

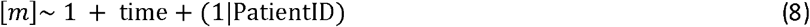

This model assumes a fixed slope (d[m]/dt) across patients, with age, gender, and PatientID contributing variable intercepts.

And Model 3:

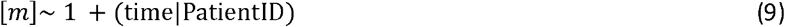

where time is included as a random slope by patient, allowing each patient to have their own trajectory of metabolic change. Likelihood ratio tests(94) were used to assess whether including random slopes significantly improved model fit.

## 4 Results

### 4.1 Phantom Experiments

#### 4.1.1 B_1_ ^+^maps

Figure 1a-b shows the reference phantom used for the acquisition of the B_1_ maps and for the validation of the absolute quantification methods. Figure 1e shows a mid-transverse slice from the B_1_^+^ map. This shows a strong transmit field close to the edges of the phantom and a B_1_^+^ dropout in the right-anterior region. The average B_1_^+^ for the mid-transverse slice shown in Figure 1e is 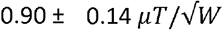 (mean ± SD, with range: 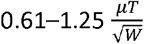). Across the whole phantom it is 0.82 ± 0.19 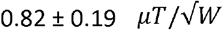 (range: 0.14–1.26 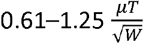 ).

#### 4.1.2 Validation of the absolute quantification method

For the reference phantom, a *T*_1_ HDO of 697 ms was measured. For the multi-compartment phantom, *T*_1_ Lac = 251 ms, *T*_1_ Fum = 314 ms and *T*_1_ HDO = 567 ms.

Results validating the absolute quantification method are shown in Figure 3. For the calibration scan, the non-calibrated HDO amplitude map is shown in Figure 3a, with saturation-corrected and calibrated maps in Figures 3b–c. As expected, calibration yielded a uniform concentration matching exactly the phantom value. For the repeat scan, the calibrated [HDO] map (as per Equation 5, Figure 3d) gave an average concentration of 1.00 ± 0.05 mol/L (range 0.87–1.25 mol/L), a 12% error relative to the true value (1.14 mol/L).

**Figure 3:**
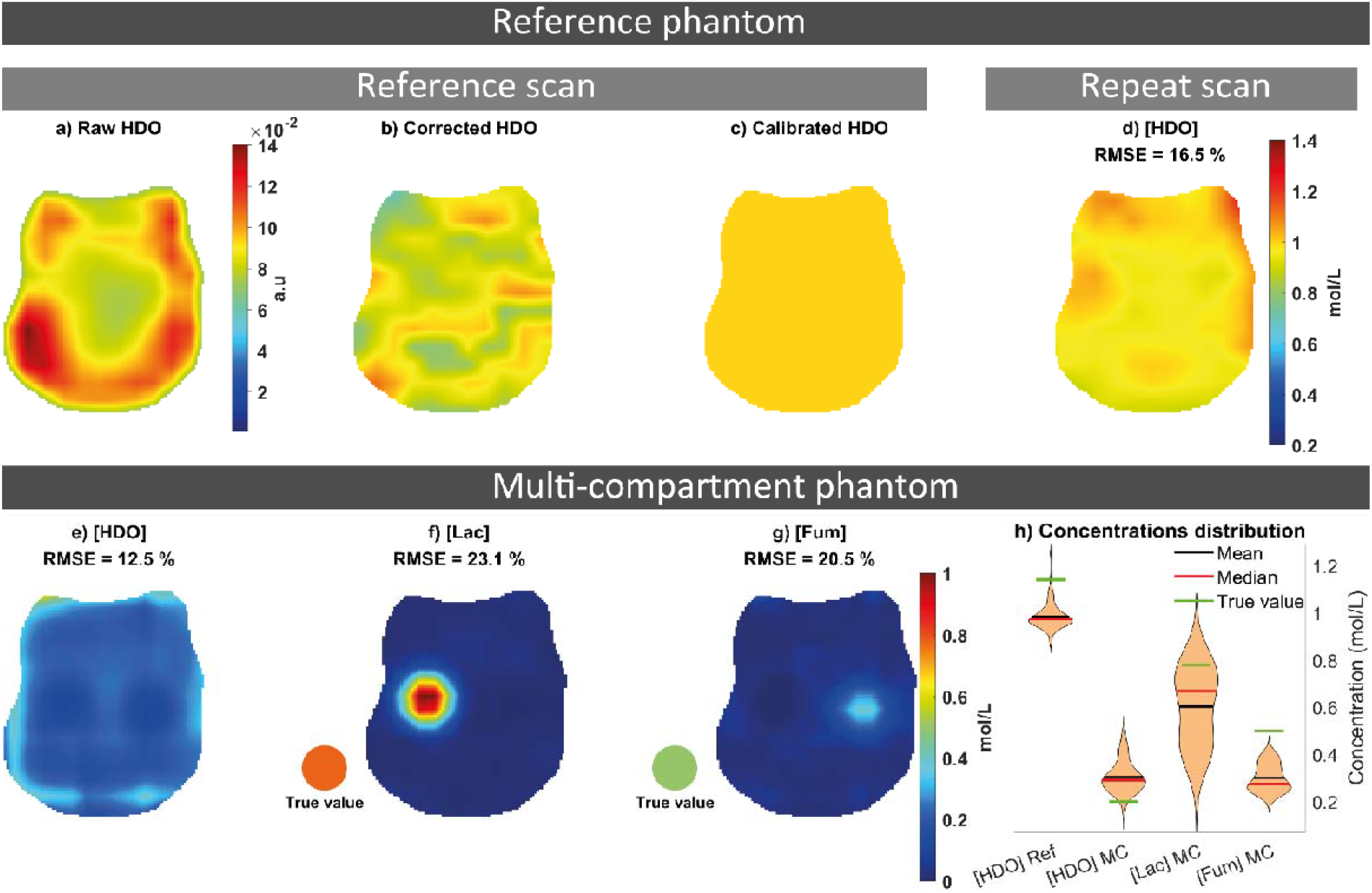
Phantom validation of absolute quantification concentration measurements. (a-d) reference phantom containing 1.14 mol/L HDO. a) Raw HDO signal in the calibration scan without calibration for absolute quantification. b) Corrected HDO signal for partial saturation in the calibration scan using Equation (5). c) quantitative HDO map after calibration for absolute quantification in the calibration scan. d) Quantitative HDO map after calibration for absolute quantification using a CSI scan acquired on a different date (repeat scan). Phantom validation of concentration measurements on the (e-h) multi-compartment phantom containing 0.2 M HDO deuterons, 0.78 M Lac deuterons (in left ball) and, 0.5M Fum deuterons (in right ball). [HDO], [Lac] and [Fum] metabolite maps are shown in e), f) and g), respectively after calibration using Equation (5). h) Distribution of the [HDO] values within the reference phantom, and of the [HDO], [Lac] and [Fum] within each whole 3D compartment of the multi-compartment (MC) phantom.

In the multi-compartment phantom (Figures 3e–g), average concentrations were: [HDO] 0.30 ± 0.07 mol/L (range 0.14–0.50 mol/L, 33% error), [Lac] 0.67 ± 0.18 mol/L (range 0.47–0.89 mol/L, 14% error), and [Fum] 0.30 ± 0.06 mol/L (range 0.25–0.39 mol/L, 40% error). Figure 3h shows the distribution of [HDO], [Lac], and [Fum] within each compartment of each phantom.

### 4.2 In vivo DMI

#### 4.2.1 Natural Abundance scans in Healthy Volunteers

Sample HDO maps from natural abundance scans in one healthy volunteer are shown in Figure 4. Mean Linewidths across volunteers varied from 11 to 26Hz in the brain. The strong signal peaks in posterior brain due to high receive sensitivity as seen in Figure 4c are corrected yielding a much more biologically plausible concentration distribution (Figure 4d).

**Figure 4:**
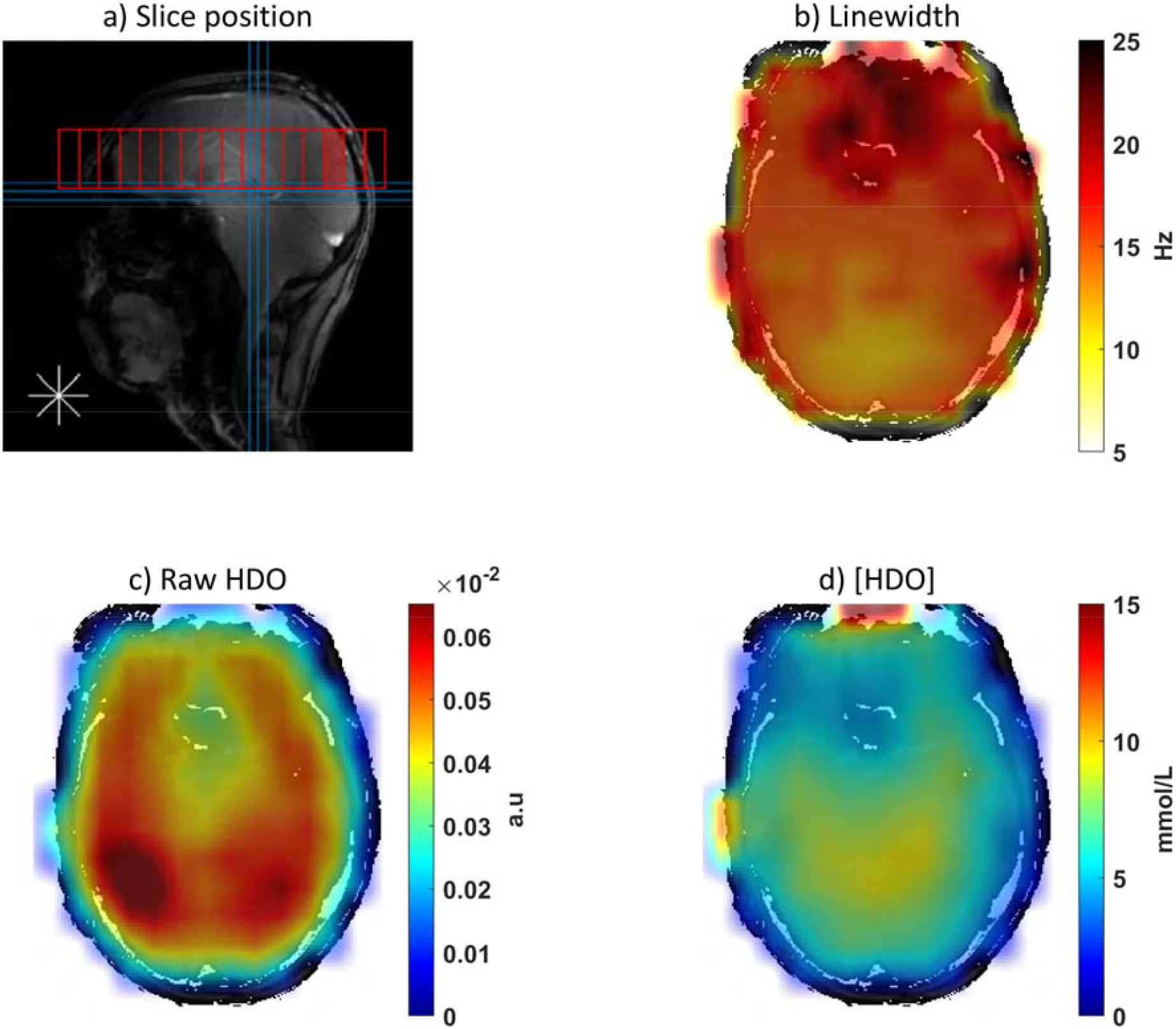
Natural HDO abundance scan in a healthy volunteer. a) Position of the transverse slice. b) Linewidth (FWHM) map, illustrating increased linewidth in the frontal lobe near the sinuses. c) Raw HDO map (i.e. not calibrated, prior to absolute quantification). d) Calibrated map showing absolute [HDO].

Figure 5 shows natural [HDO] maps for all volunteers, with violin plots depicting the distribution of [HDO] across the brain. The average HDO concentration in the brain in the 10 healthy volunteers was 8.96 ± 0.7 mmol/L (range 7.5–10.1 mmol/L).

**Figure 5:**
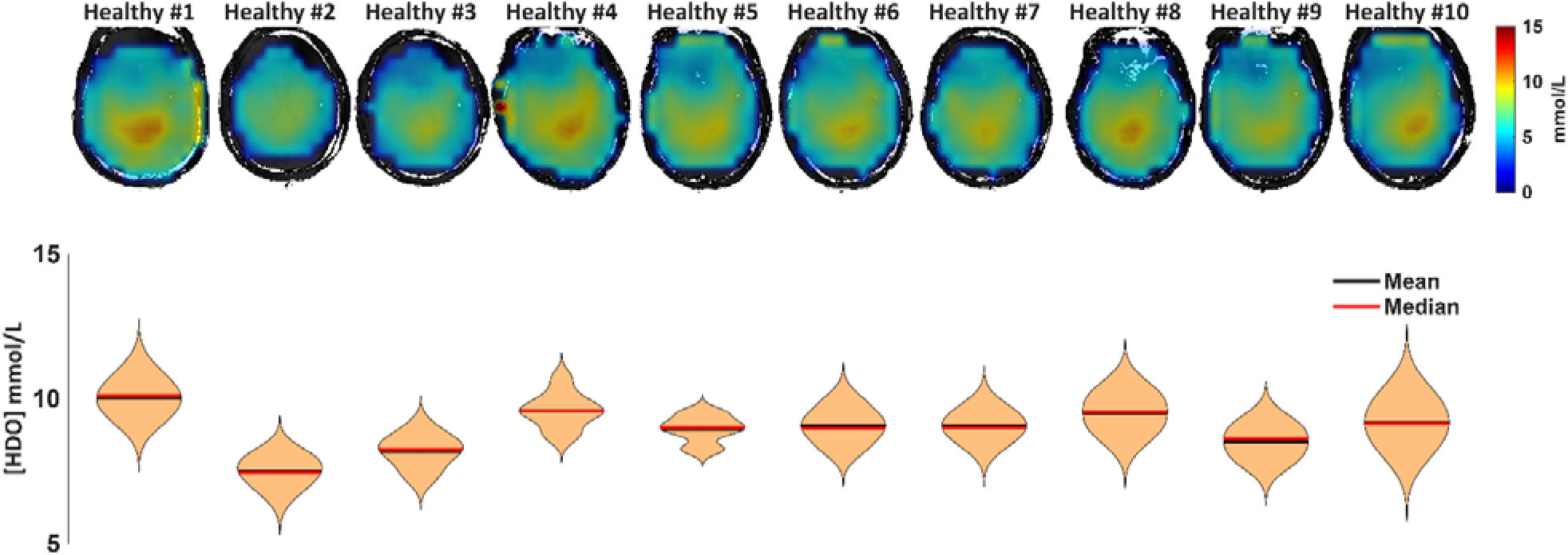
[HDO] maps for all healthy volunteers scanned with natural abundance (1^st^ row). Distribution of the [HDO] values within the whole 3D brain (2^nd^ row) shown as violin plots.

#### 4.2.2 Imaging glucose metabolism in healthy controls

^2^H images from a healthy volunteer after oral [6,6-^2^H_2_]-glucose are shown in Figure 6. Maps of HDO (Figure 6a), Glc (Figure 6b), Glx (Figure 6c), and Lac (Figure 6d) over time are presented. Signals from HDO, Glc, and Glx increased, with higher HDO in the brain center. As in prior DMI studies, a peripheral “Lac” ring is visible, which is said to be an artefact from subcutaneous lipids(96) .

**Figure 6:**
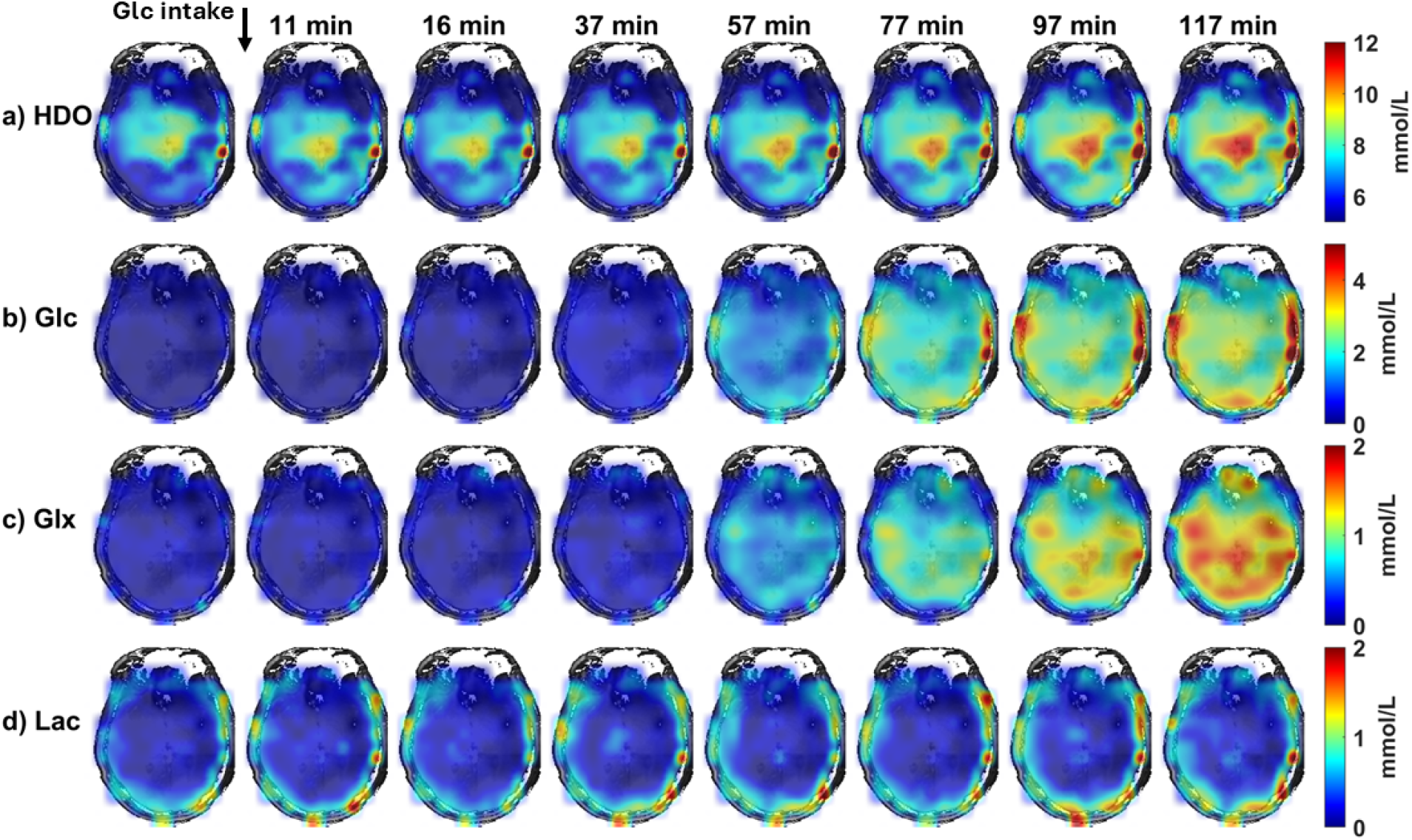
Evolution of metabolite maps evolution following oral [6,6-^2^H_2_]-glucose administration to a healthy volunteer. HDO, Glc, Glx and Lac maps are shown in a), b), c) and d), respectively after application of the whole brain mask #1.

#### 4.2.3 Imaging glucose metabolism in patients with glioblastoma

Images from glioblastoma patients are presented in Figures 7-10. Example spectra from a tumor region and a control region are shown in Figure 7b-i. Deuterated metabolite time courses for the respective voxels are shown in Figure 7j-m. Increasing HDO, Glc and Glx concentrations were observed following the administration of labelled glucose. A higher Lac concentration uptake in the tumor can be observed in Figure 7m and an opposite trend can be seen for the Glx concentration (Figure 7l).

**Figure 7:**
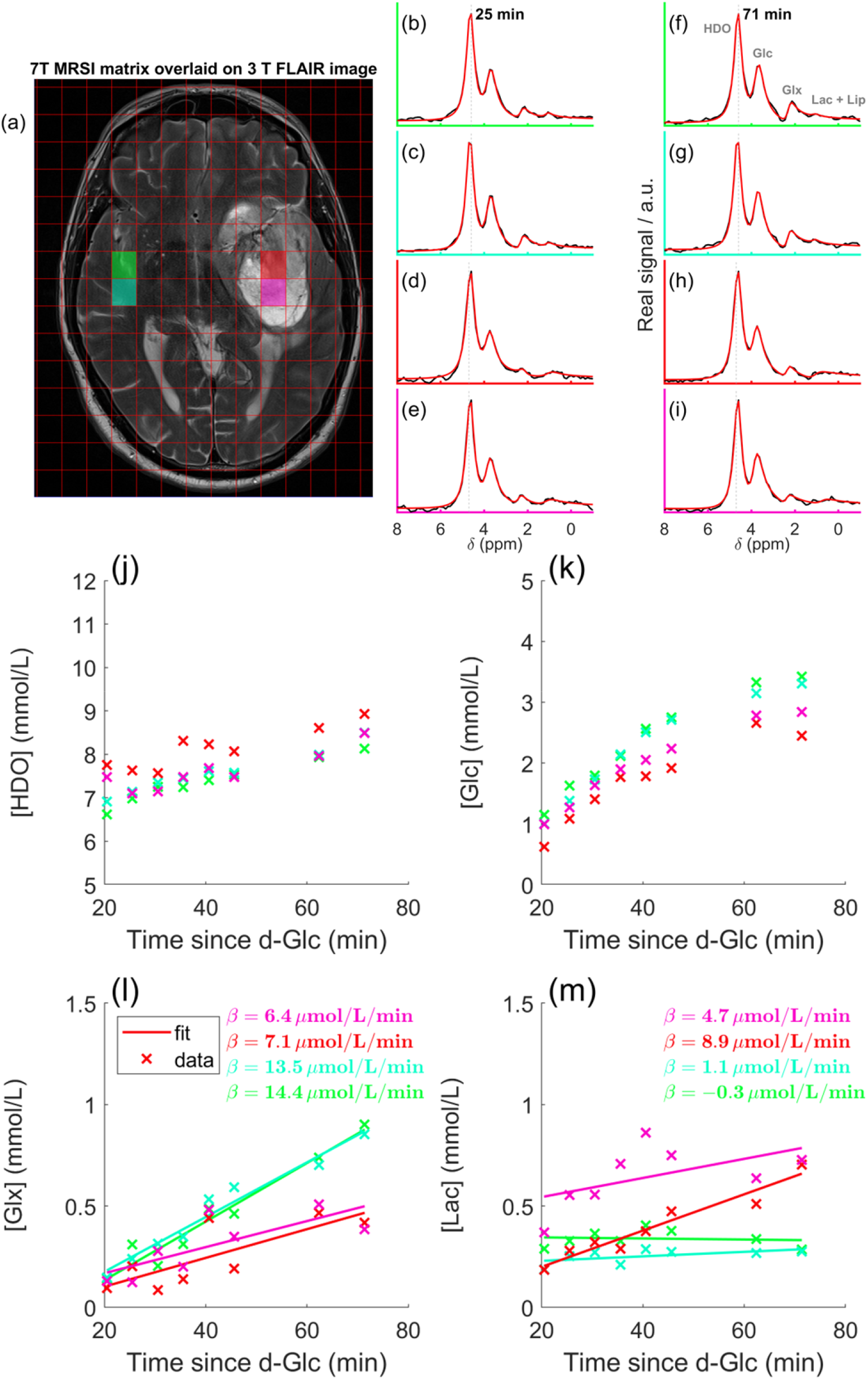
^2^H-MRS data from a glioblastoma patient. Spectra for 2 different time points from the highlighted voxels in control (green, light-blue) and tumour (red, pink) regions. Spectra from 25 min (b-e) and 71 min (f-i)) are shown from the highlighted voxels in a), overlaid on an anatomical 3T image. Time courses for each metabolite are shown in (i-m) following Glc administration. Linear fits according to Equation (7) are shown in l) and m).

Figure 8 shows the Glx concentration maps (row 1) derived from the last 3 time points for one healthy volunteer and the five glioblastoma patients. Also shown is the rate of increase in the product selectivity towards Lac ratio (row 2). A lower Glx signal and a higher ratio was observed in the tumor when compared to normal appearing brain tissue. Glc and Lac concentration maps are also shown in Figure SI3. Figure SI4 illustrates the benefits of applying an additional mask (mask #2), which excludes voxels with CRLB values of the ratio greater than 30%.

**Figure 8:**
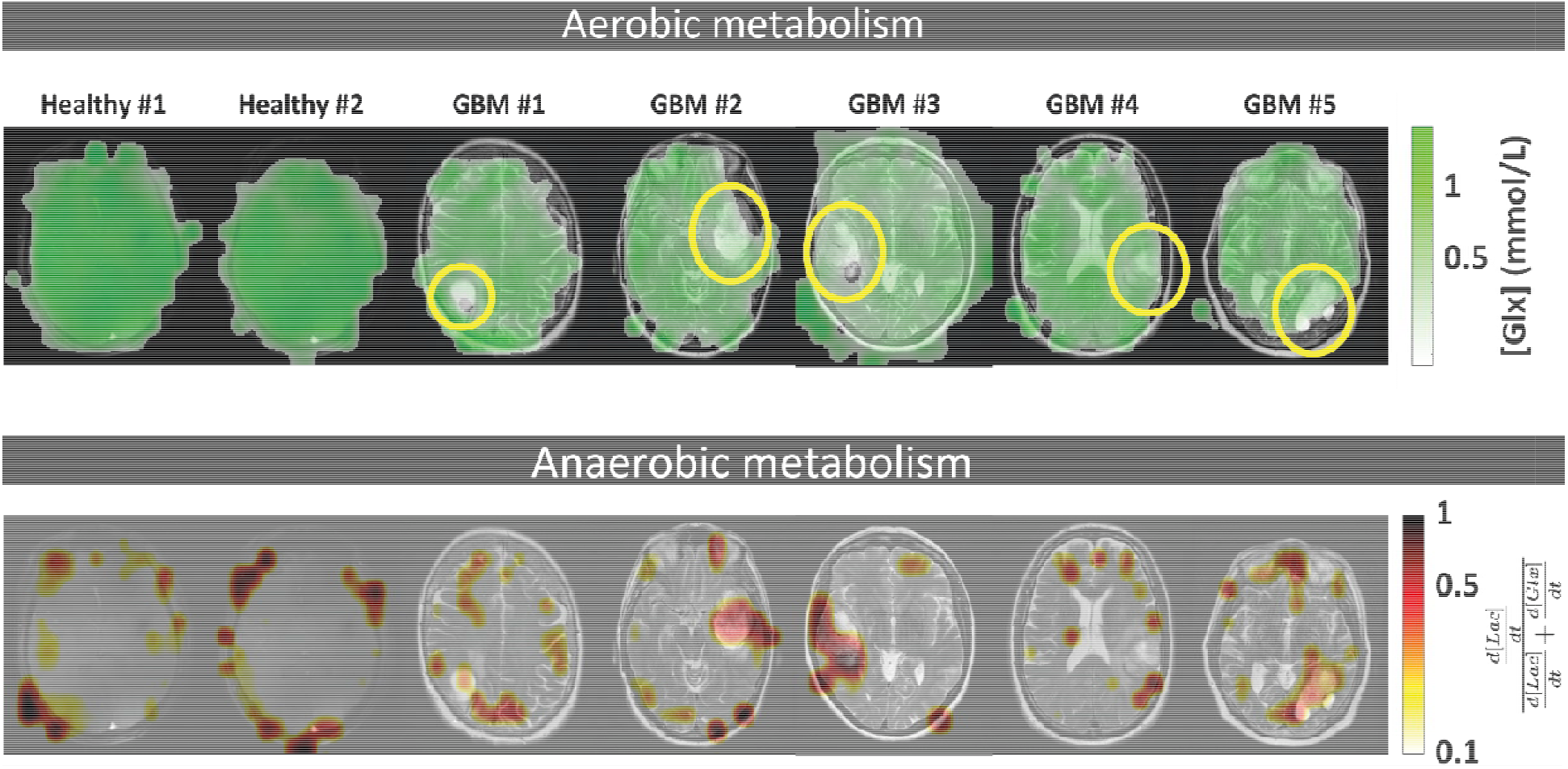
Maps of [Glx] (mmol/L) (top row, based on mask ) and the *S*_Lac_ ratio (bottom row) for the two healthy volunteers and the 5 glioblastoma patients (based on mask #2). Tumor regions are circled in light yellow.

These findings are further supported by averaged voxel data from healthy volunteers (ROI_HV_), and glioblastoma patients in control (ROI_NAWM_) and tumor (ROI_Tumor_) regions (Figure 9a–b).

**Figure 9:**
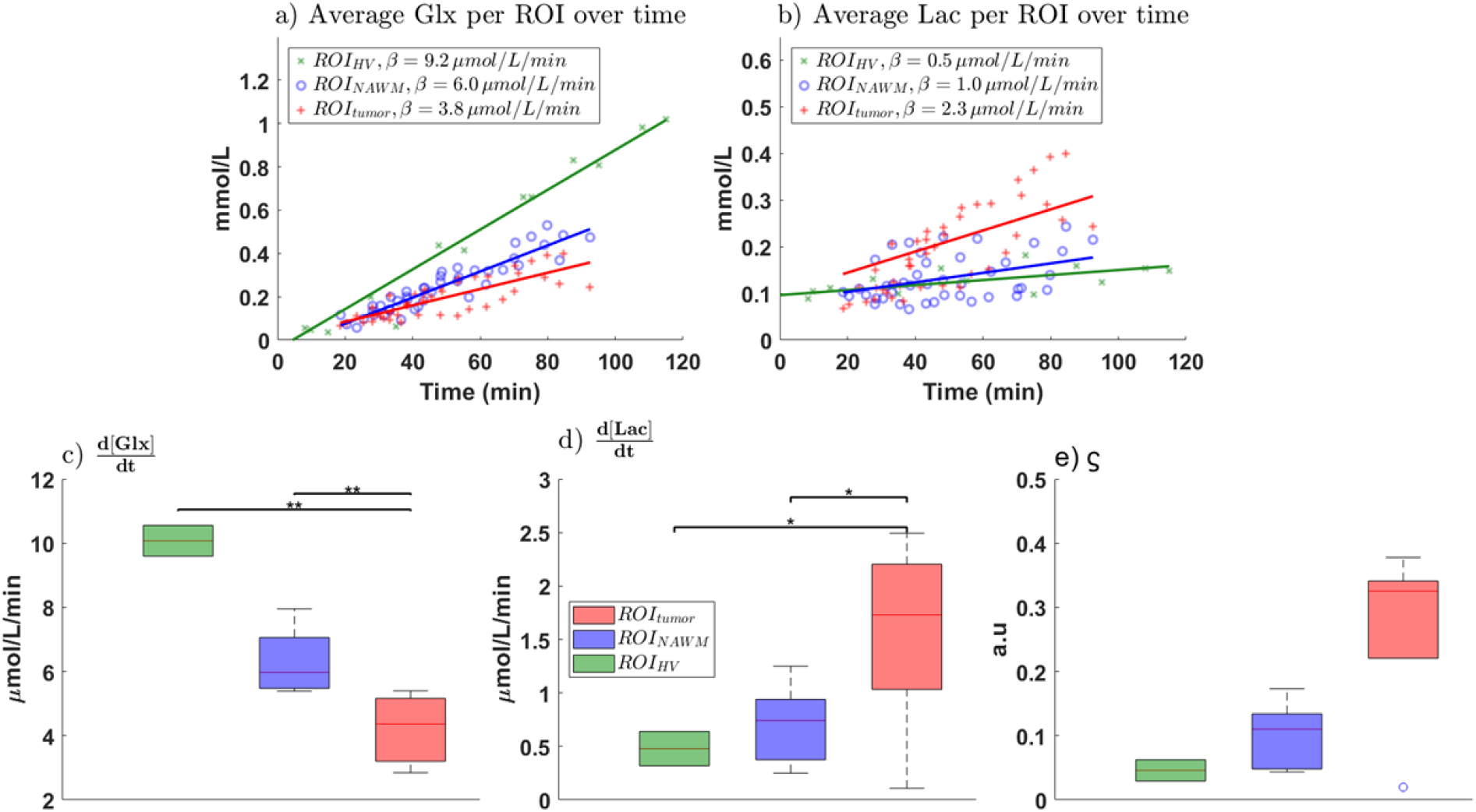
Comparison of Glx and Lac production in healthy volunteers and glioblastoma patients based the different ROis. (a) Glx and (b) Lac concentrations. Each data point represents an individual measurement per region (ROI), per time point (section 3.7.2) from a volunteer or patient, and all data within each group/region were fitted with a linear model (healthy volunteers, ROI_HV_ = green; glioblastoma normal-appearing white matter, ROI_NAWM_ = blue; tumor region, ROI_Tumor_ = red). Linear fits are also shown for each region. Rates of Glx (c), Lac (d) and *S*_Lac_ (e) production in each region (* = significant difference, p<0.01, ** = highly significant, p<0.001).

For Glx, the fitted rate of increase (d[Glx]/dt) differed significantly between tissue types. Tumor regions showed lower production rates (3.8 μmol/L/min, SE = 0.44; 95% CI: 2.88-4.66) compared with NAWM (6.0 μmol/L/min, SE = 0.36; 95% CI: 5.29-6.75; _p_ < 0.001) and healthy volunteers (9.2 μmol/L/min, SE = 0.61; 95% CI: 7.86-10.53; _p_ < 0.001).

For Lac, the fitted rate of increase (d[Lac]/dt) was significantly higher in tumors (2.3 μmol/L/min, SE = 0.87; 95% CI: 0.50-4.02) compared with NAWM (1.0 μmol/L/min, SE = 0.36; 95% CI: 0.28-1.74; _p_ < 0.01) and healthy volunteers (0.5 μmol/L/min, SE = 0.17; 95% CI: 0.17-0.91; _p_ < 0.01).

Overall, Glx production rates decrease from healthy tissue to NAWM and further to tumors (Figure 9c), whereas Lac production shows the inverse pattern (Figure 9d). The ratio similarly increases in tumor tissue (Figure 9e).

#### 4.2.4 Metabolic heterogeneity

Pairwise comparisons of tumor (ROI_Tumor_) and normal-appearing regions (ROI_NAWM_) revealed significant differences in both Glx and Lac (Glx: p<0.001, z = 4.6, r = 0.7; Lac: p<0.001, z = -4.4, r = - 0.67). Mean regional differences were -0.06 mmol/L for Glx and +0.08 mmol/L for Lac.

The best models selected by a stepwise linear modelling approach were:

with Region emerging as a significant predictor for both metabolites. For Glx, Region was associated with lower concentrations in tumors (p<0.001, β = -0.06 ± 0.01 mmol/L, 95% CI: -0.07 to -0.04; R^2^ = 0.876). For Lac, tumors showed higher concentrations (p<0.001, β = 0.08 ± 0.01 mmol/L, 95% CI: 0.05–0.10; R^2^ = 0.759) (Figures SI5–SI6, Fig. 10k–l).

**Figure 10:**
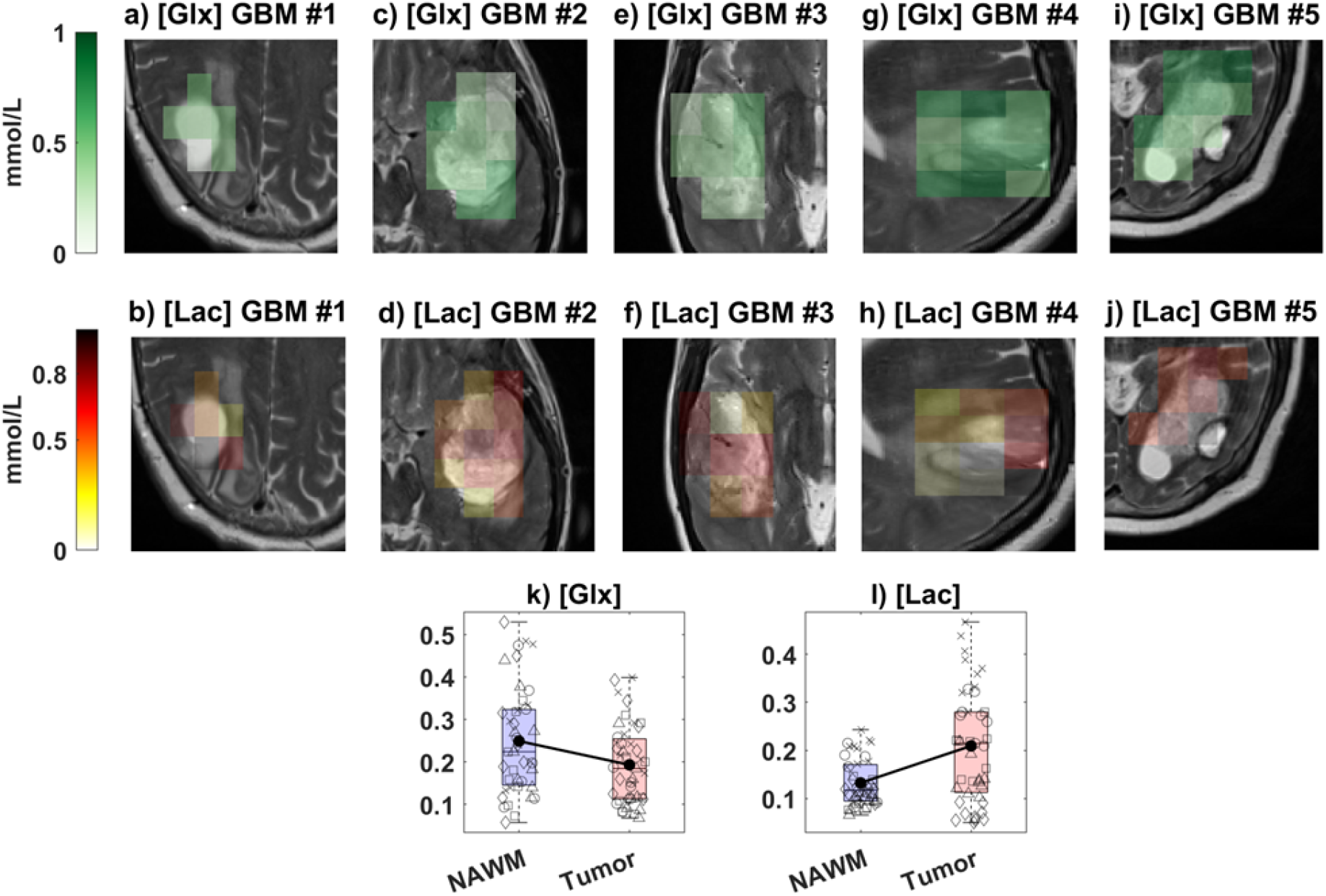
Metabolic heterogeneity in glioblastoma. [Glx] (a, c, e, g, i) and [Lac] (b, d, f, h, j) maps in all glioblastoma patients at the last time point. Boxplots showing concentrations of (k) Glx and (l) Lac for all 5 patients in NAWM and tumor regions. Black lines connect group means. For Glx, linear mixed modelling showed a significant effect of Region (β = –0.056, SE = 0.009, 95% CI [–0.073, –0.039], p < 0.001). For Lac, concentrations were significantly higher in tumor regions (β = 0.077, SE = 0.011, 95% CI [0.056, 0.098], p < 0.001). Each data point used for the boxplots corresponds to the average concentration within all voxels of the ROI (tumor or NAWM) for a given patient at a given time point, i.e. one data point per region per patient per time point.

A linear mixed-effects model (Model 1, Eq. 10) further confirmed Region as a significant determinant, with random intercepts and slopes by patient (Glx: F(1,84)=49.5, p<0.001, β=-0.06; Lac: F(1,84)=53.8, p<0.001, β=+0.08).

Figures 10a–j illustrate spatial heterogeneity in three representative patients. Across all patients at the last time point, mean tumor concentrations were 0.5 ± 0.2 mmol/L for [Glx] (0.02–0.9 mmol/L range) and 0.5 ± 0.2 mmol/L for [Lac] (0.1–1.2 mmol/L range).

Likelihood ratio tests indicated that adding time as a random effect significantly improved model fit over random-intercept-only models (Model 2 vs Model3, Eq. 8 vs. Eq. 9). For Glx, random slopes yielded a better fit (p<0.01, LR = 12.8), and for Lac, a similar improvement was observed (p<0.01, LR = 4.6).

Accordingly, linear mixed-effects models with random intercepts and slopes for time (Model 3, Eq. 9) were fitted separately for Glx and Lac using tumor-only data. These models included time as a fixed effect while capturing inter-patient variability in both baseline levels and temporal trajectories.

For Glx, there was significant variability in intercepts (SD = 0.061, 95% CI: 0.026–0.145) and slopes (SD = 0.00098, 95% CI: 0.00046–0.00208). A strong negative intercept–slope correlation (r = –0.996) indicated that patients with higher baseline Glx tended to show slower—or even negative— temporal changes.

For Lac, variability was also evident in intercepts (SD = 0.077) and slopes (SD = 0.00069), though confidence intervals could not be reliably estimated due to limited patient numbers. Nevertheless, the model suggests meaningful differences in lactate dynamics across patients.

Together, these results confirm that there can be significantly different metabolic trajectories within tumor regions between individual glioblastoma patients, which is expected given the underlying biology of these tumors.

## 5 Discussion

### 5.1 Performance of the coil array

We demonstrated the feasibility of using a novel TEM head array coil for brain ^2^H-MRSI at 7T. B_1_^+^ maps acquired on a head-shaped phantom showed acceptable coverage across the brain. The modest anterior-right B_1_^+^ drop-out did not impede absolute quantification analyses.

Transmit efficiency was 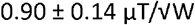 (range 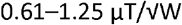 in the mid-transverse slice; 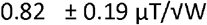, range 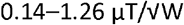 across the whole phantom), comparable to values from previous 7T DMI studies (around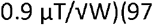). Higher transmit efficiencies have been reported in other DMI setups, such as around 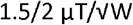 at 7T(39,93) and 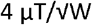 at 9.4T(98).

A key advantage of this design is its tolerance of a “missing rung,” yielding an open geometry that improves patient comfort and will enable future visual-stimulus fMRS studies. It also enables interleaved ^1^H/^2^H acquisitions(45,99,100). This provides the opportunity to implement motion correction, frequency shot-to-shot correction, and interleaved B_0_ shimming(101,102). The main limitation is B_1_^+^ inhomogeneity in frontal regions, particularly left–right asymmetry.

### 5.2 Natural abundance HDO in healthy volunteers

The absolute concentration of HDO measured in the brain of 10 volunteers (8.96 ± 0.7 mmol/L) was lower than has been reported previously (10.12 to 17.8 mmol/L(34,93,100,103)). However, to our knowledge, ours is the first study to report a directly calculated natural abundance HDO concentration in the human brain *in vivo*. Previous studies instead estimated brain HDO concentration based on the expected fraction of deuterated water in water (0.0115%(104) to 0.015%(105)) based on ^2^H natural abundance, combining this with assumed brain water fractions (ranging from 75% to 100%). Yet the fraction of deuterated water in water can vary depending on the environment(105,106) and it is well established that the water content in brain tissue varies between approximately 70–80% in gray and white matter(107,108). Based on these values, the natural HDO concentration in the brain is estimated to range between 8.9 mmol/L (assuming 0.0115% deuterated water and 70% brain water fraction) and 13.3 mmol/L (assuming 0.015% deuterated water and 80% brain water fraction). The mean value measured in this study falls within this range.

Absolute quantification of metabolites provides direct concentration values, unlike metabolite ratios, which rely on reference metabolites that may themselves vary depending on the biochemical state of the tissue. This is particularly important in heterogeneous diseases like glioblastoma, where reference metabolites can be altered by tumor metabolism, edema, or treatment(62,109) as we demonstrated by the changes in “normal-appearing” tissue in Figure 8.

Moreover, absolute quantification avoids reliance on fixed assumptions, for example an assumption that the natural abundance HDO concentration is the same in all patients. Such assumptions fail to account for inter-subject variability due to differences in hydration status, pathology, or metabolic state.

### 5.3 In vivo glucose metabolism

A linear model was fitted to the Glx and Lac concentration time courses, consistent with approaches used in other studies(40,93). We chose this simpler model because Glx and Lac concentrations did not reach a steady state; however, more sophisticated kinetic models are available for fitting concentration dynamics over time(110). The increase in HDO concentration observed in the center of the brain could be due to accumulation of HDO produced from Glc metabolism in cerebrospinal fluid (CSF has 99% water, which is higher than in the white and grey matters).

A UTE-CSI sequence(111,112) was used here that had been optimized for DMI acquisition. However, newer sequences such as SSFP(113) or EPSI(114) may improve the signal-to-noise ratio and therefore the metabolic information attainable. The spatial resolution reported here falls between those reported in recent *in vivo* brain DMI studies at 7T (2–3 cm^3^) and 9.4T (1–2 cm^3^)(31,32,39,96,97,115).

### 5.4 Assessment of tumor metabolism

In 4 of 5 glioblastoma patients, tumors showed an increased S_Lac_ ratio when compared to normal appearing brain regions, indicating increased glycolytic metabolism, as has been observed in preclinical(110) and clinical studies(92). Rates of Lac production varied between patients, consistent with preclinical studies showing the presence of different metabolic subtypes of glioblastoma with differences in glycolytic and oxidative activities(92,110) and clinical studies showing that the tumors of glioblastoma patients can show differences in the proportion of these different metabolic subtypes(62). Similar inter-patient variation has also been observed in hyperpolarized [1-^13^C]-pyruvate studies, where tumor lactate labelling differed markedly(116).

The mean [Glx] increase rate 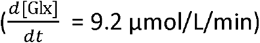 in healthy volunteers is lower than the rate of 21 μmol/L/min reported in a recent study(40), but is comparable to other preclinical studies(62,110).

Prior studies have suggested that [Glx] declines (5 – 15 % per decade) with aging(117-119), which could partially contribute to the difference in concentrations seen between healthy volunteers (24-28 years old) and glioblastoma patients (> 50 years old).

Intra-tumoral variations in [Glx] and [Lac] were also visible in all representative patients. Elevated lactate or high pyruvate to lactate conversion has been observed in hypoxic, poorly perfused regions (tumor rim *vs* core)(116), while [Glx] changes are often found at invasive margins, with the direction of change depending on tumour subtype, local neuron–astrocyte metabolism and partial-volume effects(120-122). Moreover, intratumoral [Glx] heterogeneity appears to be associated with more aggressive tumor behavior, as seen in a recent study(122). Heterogeneous glucose turnover has been observed previously where some tumor regions have higher glycolytic flux while others rely more on oxidative metabolism (OXPHOS)(110,123). For this reason, achieving higher spatial resolution in DMI would be valuable, and might be facilitated by advanced analysis methods such as deep learning, low-rank approaches, or model-based reconstruction techniques(124,125).

Most glioblastoma studies have focused on [Lac] or [Lac]/[X] ratios. But these ratios are difficult to interpret when both numerator and denominator approach zero, often yielding misleading values. Our use of absolute quantification enables robust detection of [Glx] differences, highlighting [Glx] as a potentially more reliable biomarker of tumor heterogeneity in DMI. Future work should emphasize quantitative [Glx] analysis and consider full kinetic modelling to better capture fluxes, although this lies beyond the scope of the present study.

These metabolic heterogeneities in between tumours and patients could be due to the tumour microenvironment (TME), cellular composition, genetics or be driven by therapy effects(20,22,126).

The ability to assess these metabolic phenotypes non-invasively *in vivo* with DMI could improve the precision of targeted therapies(62,127).

A limitation of our study is the small sample size, nevertheless the observation of intra- and inter-tumor heterogeneity warrants larger-scale studies.

## 6 Conclusion

We present step-by-step a method for absolute quantification of ^2^H MRS images, which we validated using a TEM head array coil on phantoms and healthy volunteers. Studies in glioblastoma patients revealed both inter- and intra-tumor metabolic heterogeneity. We anticipate that this new method will enhance our understanding of brain metabolism and tumor behavior, as well as aid in optimizing drug therapies and monitoring treatment response.

## Abbreviations

RF: RadioFrequency
FID: Free Induction Decay
CSI: Chemical Shift Imaging
DMI: Deuterium Metabolic Imaging
WSVD: Whitened Singular Value Decomposition
AMARES: Advanced Method for Accurate, Robust and Efficient Spectral fitting
AIC: Akaike Information Criterion
TEM: Transverse electromagnetic
Glx: Glutamine/glutamate
Glc: Glucose
Lac: Lactate
Fum: Fumarate
HDO: semi heavy water
SAR: Specific Absorption rate
FWHM: Frequency Width to Half Maximum

## Acknowledgements

This work was supported by the EU Horizon Europe project “MITI” #101058229. This research was supported by the NIHR Cambridge Biomedical Research Centre (NIHR203312) and the NIHR Applied Research Collaboration East of England. The views expressed are those of the author(s) and not necessarily those of the NIHR or the Department of Health and Social Care.

Minghao Zhang was funded by the Medical Research Council (MR N013433-1) and the Cambridge Trust. Masha Novoselova was funded by the Commonwealth Scholarship Commission and the Cambridge Trust. Carina Graf was supported by the Cambridge Trust, the W.D. Armstrong Fund and the European Union’s H2020 research and innovation program under grant agreement [801075].

We thank the WBIC radiographers for their assistance with the GBM patient scans. We thank Prof Carpenter and Prof Williams for reviewing our coil safety documentation.

We thank the Department of Radiology and Cambridge University Hospitals NHS Trust Pharmacy for assistance storing the 6,6’-[ ^2^H_2_]-glucose appropriately and for defining the procedure for administration.

## Data availability

The code is hosted on GitLab and can be made available upon request to the authors. Data is available from the corresponding author subject to a data transfer agreement as mandated by our ethical approvals and hospital policies.

## 7 Supplementary information

**Figure SI1:**
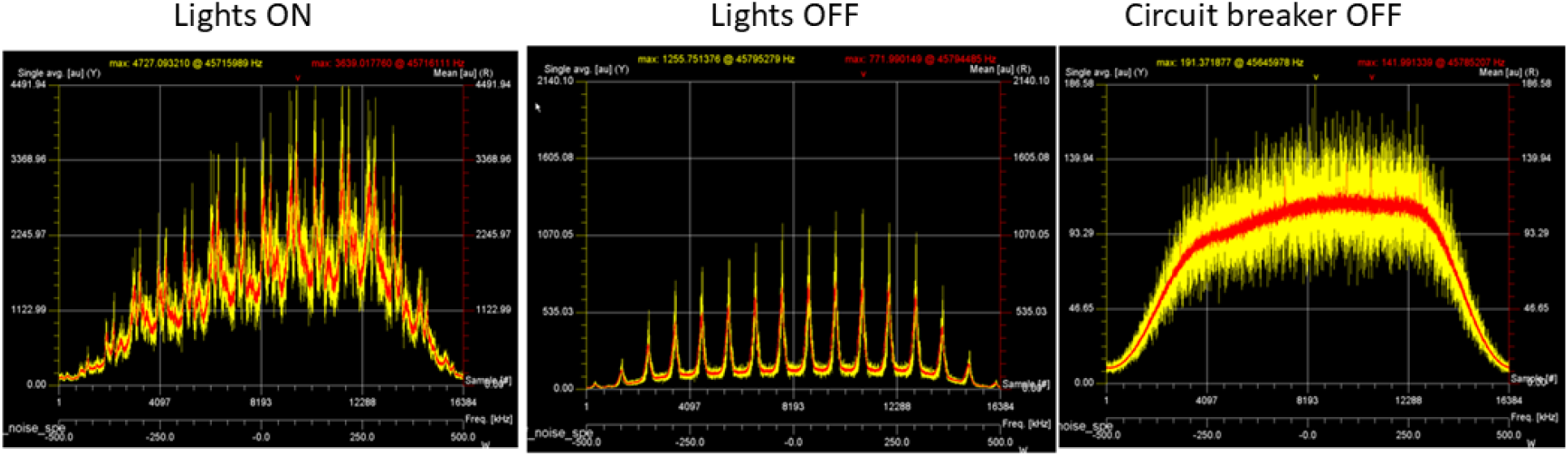
Effects of the lights on the environment noise captured with the coil’s receive channels. Difference in noise level between lights ON, lights OFF and circuit breaker turned OFF completely in the 7T “technical room”.

**Figure SI2:**
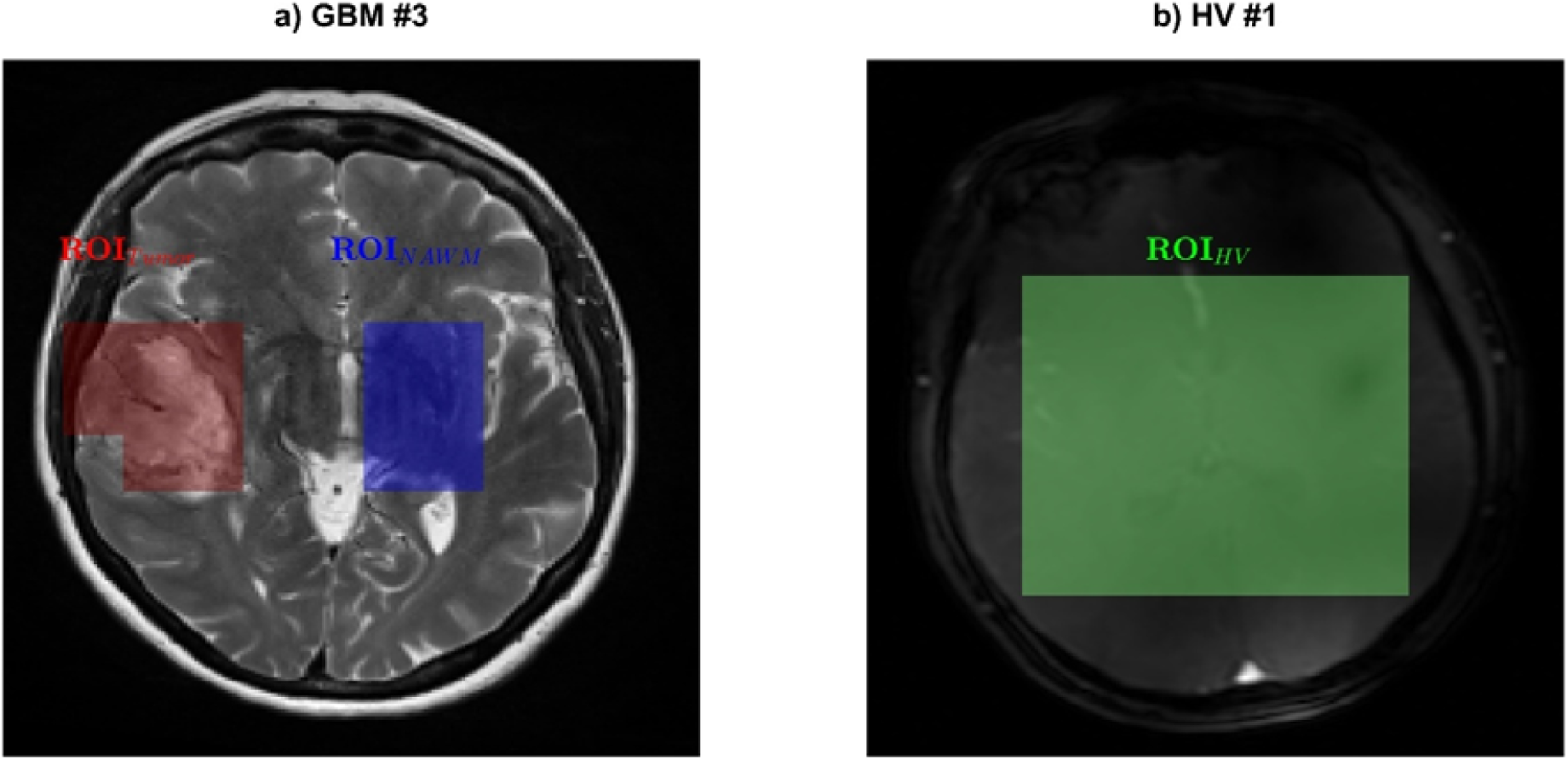
Illustration of the different ROIs that have been chosen for further statistical analysis on a) a glioblastoma patient (ROITumor and ROINAWM) and b) a healthy volunteer (ROIHV).

**Figure SI3:**
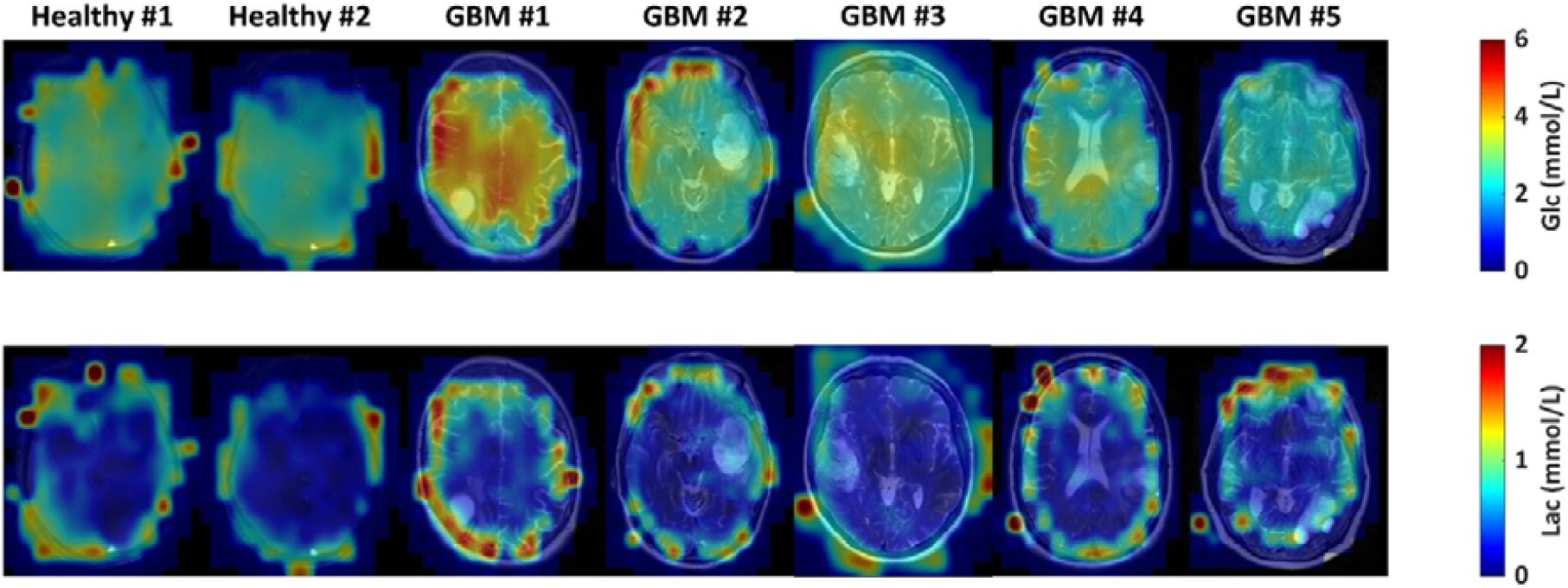
Maps of [Glc] (mmol/L) (top row) and [Lac] (bottom row) for the two healthy volunteers and the 5 glioblastoma patients based on mask #1 averaged across the last three time points.

**Figure SI4:**
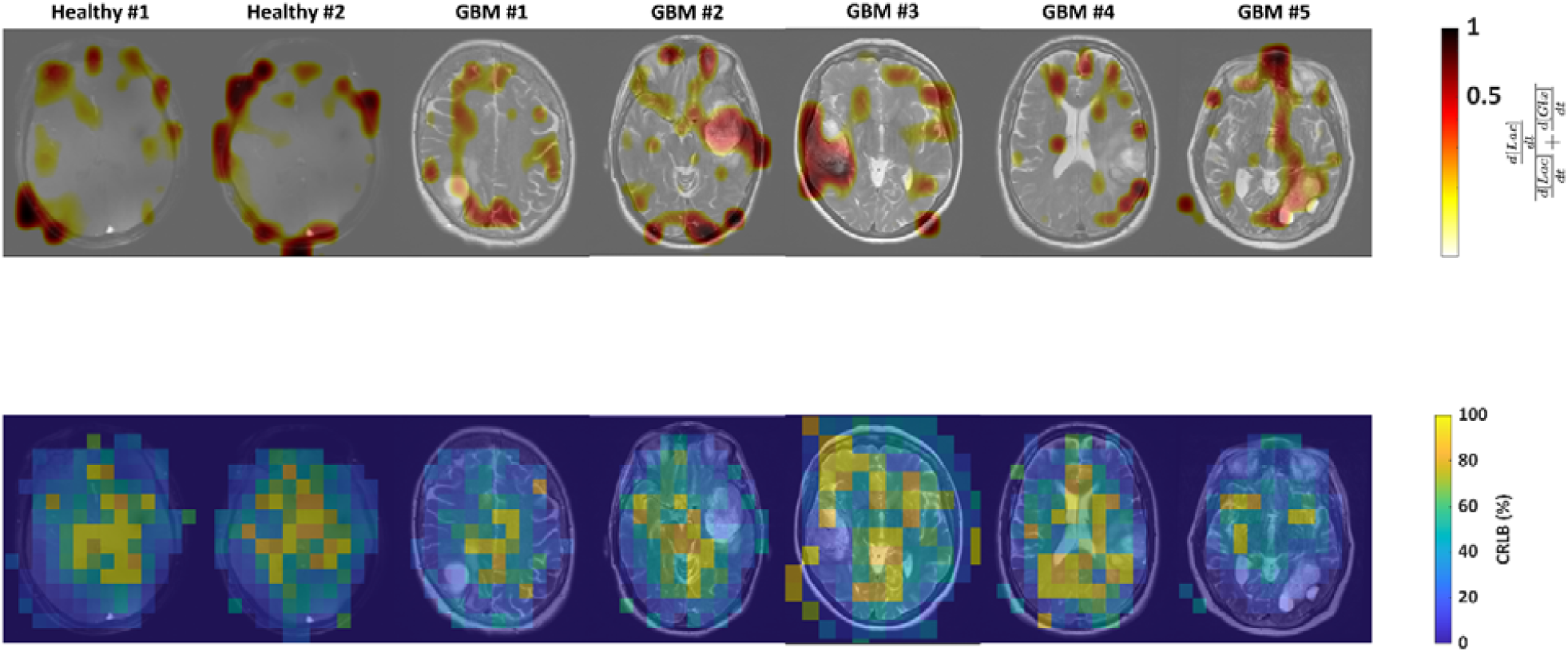
Maps of *S*_Lac_ ratio (top row) for two healthy volunteers and five glioblastoma patients, based on mask #1 (i.e., without excluding voxels with CRLB values of the ratio >30%). The bottom row shows the corresponding CRLB maps for the same subjects. Note that the % CRLB is highest in areas with low metabolic rate -- i.e. low d[Lac]/dt and low d[Glx]/dt.

**Figure SI5:**
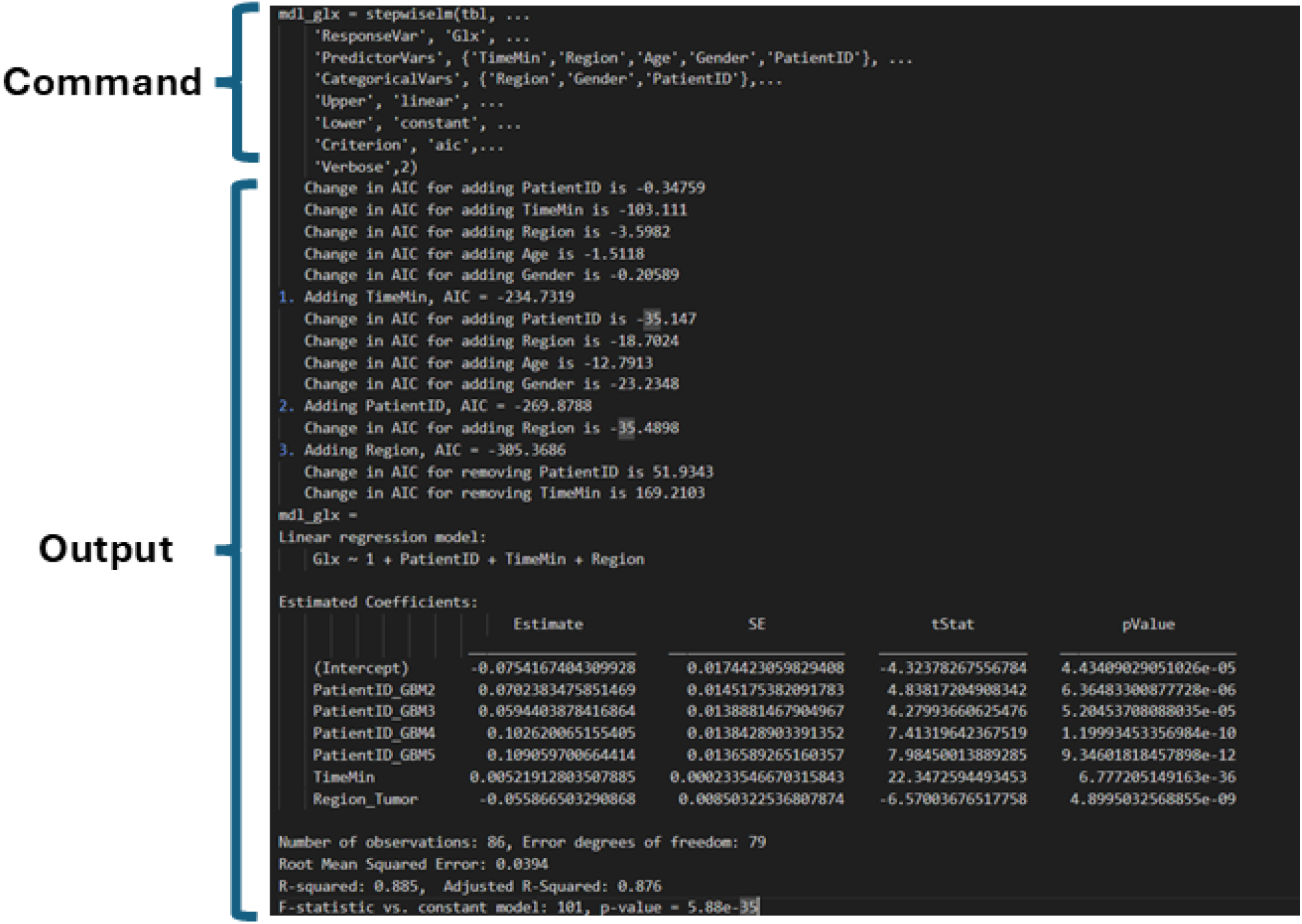
Matlab code and output from the stepwise linear modelling of [Glx] using the Akaike Information Criterion (AIC).

**Figure SI6:**
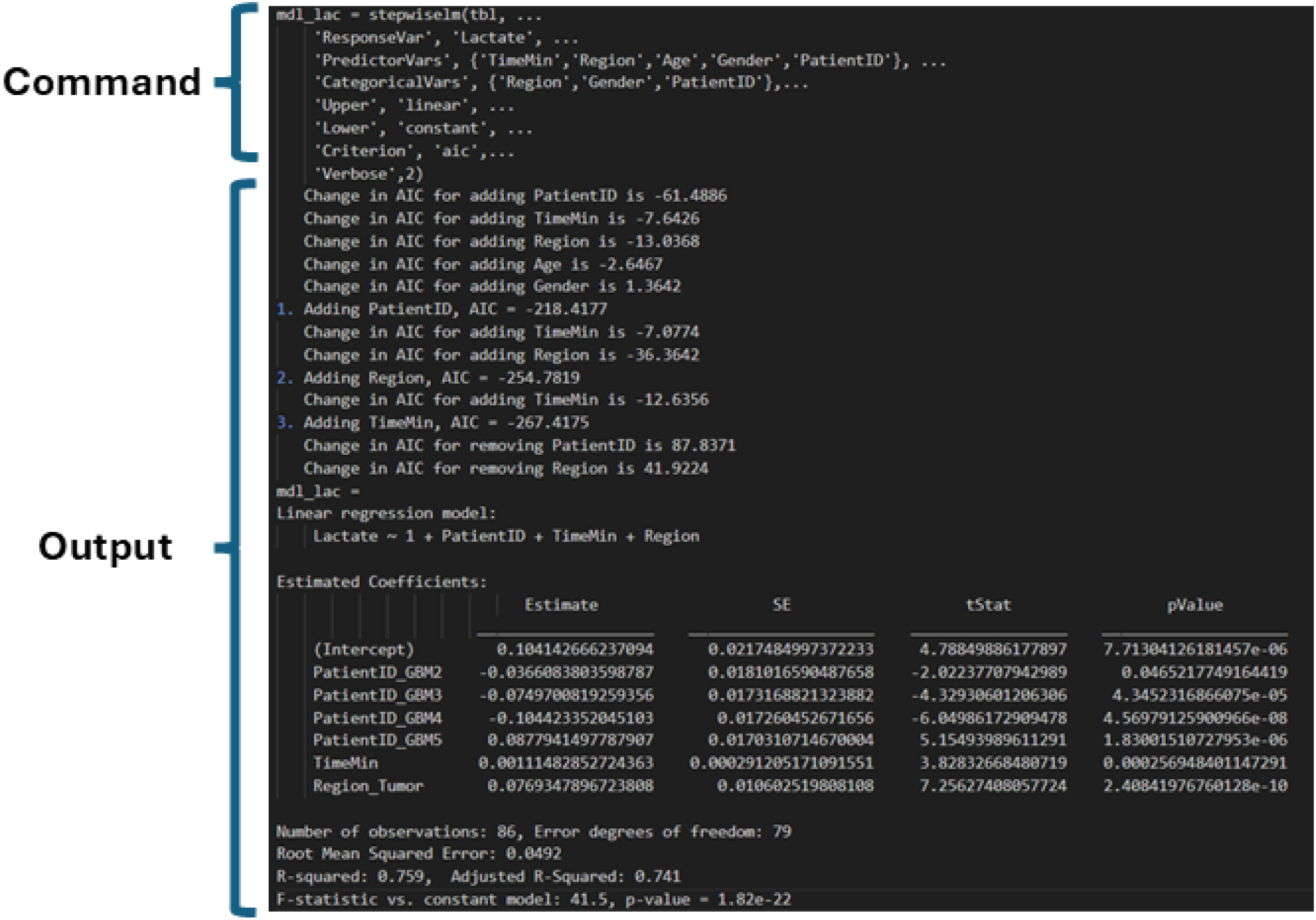
Matlab code and output from the stepwise linear modelling of [Lac] using the Akaike Information Criterion (AIC).

